# Neuroimaging Correlates of Post-Stroke Pain After Ischemic Stroke: Secondary Analysis of the INSPiRE-TMS Trial

**DOI:** 10.1101/2025.08.07.25333248

**Authors:** Alina Stegemann, Ana Sofia Rios, Ahmed Khalil, Ulrike Grittner, Uchralt Temuulen, Ramanan Ganeshan, Tim Bastian Braemswig, Andreas Horn, Thomas Ihl, Heinrich J Audebert, Anna Kufner, Matthias Endres

## Abstract

**Background:** Post-stroke pain (PSP) affects nearly half of stroke survivors, severely compromising quality of life. The causes of PSP remain underexplored, although there is likely a complex interplay of lesion effects, psychological factors and mobility that play a role in the development of PSP. The aim of the study was to investigate clinical characteristics associated with PSP, as well as structural and functional correlates of PSP using lesion symptom (LSM) and network mapping (LNM).

**Methods:** We analyzed data from the INSPiRE-TMS cohort, encompassing 1,022 minor ischemic stroke patients. Pain severity and psychological factors (EQ5D-3L questionnaire) were assessed annually for up to 3 years. In a sub-group of 391 patients with available imaging data, LSM and LNM analyses were conducted to identify neural correlates of PSP.

**Results:** Overall, 47% of patients reported pain 1-year post-stroke. Multivariable regression analyses identified baseline anxiety as being associated with PSP assessed at 1-year post-stroke (OR 2.90, 95% CI 1.17–7.17, p=0.021). LSM did not identify any voxels associated with new severe pain. LNM identified a network involving anterior cingulate cortex, thalamus and insular cortex. Adjusting for anxiety highlighted distinct network contributions, suggesting interactive effects of psychological states on pain perception. Automated comparison to large metanalytical findings using Neurosynth associated the terms ‘pain’ and ‘nociception’ most strongly to the identified network.

**Conclusion:** PSP is closely associated with psychological factors such as anxiety. LNM of PSP revealed disruptions in a pain-related neural network consistent with prior pain research. These results warrant external validation and could guide future network-targeted neuromodulation therapies.

**Registration:** URL: https://www.clinicaltrials.gov; Unique identifier: NCT01586702.

## Introduction

Stroke is a major global health concern, profoundly impacting patients’ quality of life. Among its various complications, post-stroke pain (PSP) is particularly debilitating, affecting both physical and emotional well-being. PSP has been linked to reduced mobility^1^, independence^2^, and difficulties with daily activities^3^, as well as increased anxiety, depression^2,4,5^, fatigue^6^ and overall decreased quality of life^1,5,7^. Recognizing the multifaceted nature of PSP is critical not only for accurate diagnosis but also for developing effective therapeutic strategies.

Clinical studies have found that the frequency of PSP is high, affecting up to 50% of stroke patients^3,7^. The onset of PSP can vary widely, occurring from one month up to two years post-stroke, and persisting for up to five years^1^. While PSP encompasses diverse etiologies – including spasticity, shoulder pain, central (neuropathic) pain, and headache^3,8^ – its complexity extends beyond physical causes. Female sex, older age, depression, spasticity, reduced upper extremity mobility and somatosensory impairment, among others, have been described as risk factors^8,9^. Moreover, psychological factors such as anxiety and depression may influence the experience and intensity of PSP^2,4^, a phenomenon well-documented in many chronic pain conditions^10–12^, but less so in the context of stroke.

Imaging studies suggest that lesion location – such as within the ascending somatosensory pathways^13,14^, specifically spinothalamic tract and thalamus^15–17^, basal ganglia^16,18^, brainstem^16,17,19^, midbrain^20^, insular and somatosensory cortex^19^ – play a central role in the development of pain following stroke. This is particularly well-described for central neuropathic PSP^14^.

Recent advances in neuroimaging have shifted the focus from isolated brain regions to whole-brain network analyses, using methods such as lesion network mapping (LNM)^21,22^. This technique allows for a more comprehensive understanding of how disruptions in specific networks contribute to the development of a specific symptom, such as PSP. Studies using this method have highlighted the role of networks involving the thalamus, insular-opercular cortex, and cingulate cortex in central neuropathic PSP specifically^14,15^. Moreover, the method has also shown promise in guiding targeted treatments^14^. Despite advances in LNM, studies across all PSP subtypes based on larger prospective, well-characterized ischemic stroke cohorts are still lacking.

Therefore, we set out to investigate in a comprehensive dataset of ischemic stroke patients 1) the clinical factors associated with the development of new severe pain. Furthermore, we aimed to 2) identify regions associated with the development of new and severe pain post-stroke using classical lesion symptom mapping (LSM) and to 3) identify underlying network effects associated with new and severe pain using LNM with a normative functional connectome. Ultimately, the overarching aim of the current study was to deepen our understanding of the underlying pathophysiology of PSP.

## Methods

### Study design and patient population

This study utilized data from the INSPiRE-TMS trial (NCT01586702), a multicenter, randomized trial comparing a secondary prevention support program and conventional care in patients with minor ischemic stroke or transient ischemic attack (TIA). Details of the study design and primary outcomes have been published previously^23,24^. The aim of the trial was to assess whether an intensified secondary prevention program after minor stroke and TIA could reduce the frequency of recurrent vascular events. The study population included adult patients (≥ 18 years old) with a TIA (with an ABCD2 score ≥ 3) or a minor stroke (with a modified Rankin Scale (mRS) score ≤ 2) and at least one modifiable risk factor such as hypertension, diabetes, atrial fibrillation or smoking. Exclusion criteria included malignant diseases with a life expectancy < 3 years, relevant cognitive deficits, substance dependency, or strokes/TIA caused by less common mechanisms such as dissection or vasculitis. The trial was approved by the Ethics Committee of Charité Universitätsmedizin Berlin (EA2/084/11) and by the local ethics committees at all participating centers. All patients provided written informed consent.

For this analysis, only patients with the diagnosis “minor stroke” and with information on stroke severity, as assessed by the National Institutes of Health Stroke Scale (NIHSS), were included. This study is reported in accordance with the STROBE guidelines.

### Clinical assessment and follow-up

Patients underwent clinical assessments at baseline (within 14 days of stroke onset), one year (V_1_), two years (V_2_), and three years (V_3_) post-stroke. Demographic and clinical characteristics, including age, sex, and known cerebrovascular risk factors, were documented by the primary treating physician. Stroke severity was assessed using the NIHSS. Functional outcomes were evaluated using the modified Rankin Score (mRS). Quality of life was assessed through the EuroQoL 5-Dimension 3-Level (EQ5D-3L) questionnaire, which includes dimensions for mobility, self-care, daily activities, pain/discomfort, and anxiety/depression. Each dimension is scored on a scale of 1 (no problems) to 3 (extreme problems).

The main outcome of the current secondary exploratory study was *‘new severe pain’* at V_1_ (1 year follow-up) defined by the presence of severe pain at V_1_ (EQ5D-3L = 3) but not at baseline (EQ5D-3L < 3 at baseline)^2^.

### Imaging

All patients who received an MRI scan within the acute hospital stay (within 7 days of stroke onset) underwent a standard stroke imaging protocol on a 3-Tesla Siemens scanner. Sequences included a T2*-weighted sequence, diffusion-weighted imaging (DWI), “time-of-flight” MR angiography, and a fluid-attenuated inversion recovery (FLAIR) sequence. The DWI protocol consisted of images acquired with a b-value of 1000 s/mm², non-diffusion-weighted images (b = 0 s/mm²), and an apparent diffusion coefficient map.

### Neuroimaging analysis

#### Preprocessing

Lesion delineation was manually performed by a trained rater (A.S.R.) on DWI-TRACE and FLAIR sequences and supervised by at least two experienced neurologists and/or radiology residents (A.Ku, A.Kh, R.G, B.B), all blinded to clinical data, using MRIcron (https://www.nitrc.org/projects/mricron). Further preprocessing and analyses were conducted as described previously in detail^25^. Briefly, DWI (b1000) and FLAIR lesion masks were co-registered to skull-stripped and bias-corrected DWI (b0) and FLAIR sequences by FMRIB Software Library FSL^26^ and subsequently normalized to a standard space based on the Montreal Neurological Institute (MNI152 atlas, 1 × 1 × 1 mm)^27^ using a series of linear and nonlinear registrations (SyN algorithm) in Advanced Normalization Tools in Python (ANTsPy)^28^. Following spatial normalization, a quality check was performed by overlaying the normalized lesions onto individual normalized brains to confirm proper alignment and comparing both MRI modalities (DWI and FLAIR), the normalized lesion mask with the best quality was selected for further analyses. All imaging analyses (LSM and LNM) were performed on a subgroup of patients, namely those that had available MR-imaging with lesions co-registered to MNI space as well as available pain data assessed via the EQ5D-3L at V_1_ (N = 391) (**Suppl. Figure 1**).

#### Lesion Symptom Mapping

Voxel-wise and atlas-based LSM analyses were performed using NiiStat (https://www.nitrc.org/projects/niistat/) to identify structural lesions associated with “new severe pain” at V_1_. A minimum lesion overlap of 5% was required and lesion volume was included as a covariate to account for its potential confounding effects^29^. The voxel-wise analysis involved 10,000 permutations to ensure robust statistical testing, with family-wise error (FWE) correction applied and statistical significance set at p<0.05. An atlas-based analysis was also performed using the Juelich Histological Atlas to enhance interpretability by linking findings to specific neuroanatomical regions, and to identify signals potentially too subtle to reach significance in the voxel-wise analysis^30^.

#### Lesion Network Mapping

The LNM analyses followed methods by Fox et al.^21^, and also as recently published by Rangus et al.^25^, using normalized binary lesion masks as seeds to assess functional connectivity within a normative connectome derived from 1,000 healthy fMRI scans^31^. A connectivity profile was calculated for each patient, between voxels within the lesion mask and all other brain voxels, producing Pearson correlation coefficients averaged across the 1,000 brains. Fisher z-transformed connectivity profiles were used for further analysis. Non-parametric permutation testing was performed with FSL *randomise* using a two-sample T-test to identify statistically significant lesion connections associated with “new severe pain” at V_1_. Pain scores were included in the analysis in a binarized manner. The analysis was run for 5,000 permutations for two contrasts examining connections linked to presence [1] and absence [-1] of “new severe pain” at V_1_. FWE correction to account for multiple comparisons and threshold-free cluster enhancement (TFCE) were applied. A threshold for significance was defined at pFWE < 0.05. Additionally, as part of a sensitivity analysis, anxiety was included as a covariate. Results were overlaid on structural atlases (Harvard-Oxford Cortical Structural Atlas, Harvard-Oxford Subcortical Structural Atlas, JHU White-Matter Tractography Atlas, FLIRT-normalized Cerebellar Atlas) to pinpoint connected regions and were compared with existing neuroimaging studies using the ‘neurosynth decoder’ on the Neurosynth platform^32^.

### Statistical analysis

STATA IC version 17 (StataCorp, College Station, Texas USA) was used to perform all statistical analyses. Descriptive statistics were used to summarize demographic and clinical characteristics. T-tests or Chi-square (χ²) tests were performed for group comparisons where appropriate. Bivariate and multiple logistic regression analyses were performed to investigate the associations between “new severe pain” at V_1_ and age, sex, lesion volume, NIHSS, mRS at baseline, EQ5D-3L anxiety at baseline and EQ5D-3L mobility at baseline. These variables were selected based on their clinical relevance and prior evidence linking them to pain outcomes in similar patient populations. A two-sided statistical significance level of 0.05 was considered. However, no adjustment for multiple testing was applied in this secondary exploratory study for the regression models. Interpretation of our results is based on effect size estimates and 95% confidence intervals (CI).

### Data availability statement

Data supporting the results of this study is available upon request from the corresponding author. Open source software tools were used for the pre-processing and analysis of the data, including: Lead-DBS (https://github.com/netstim/leaddbs), FSL 6.0.6.4 (https://fsl.fmrib.ox.ac.uk/fsl/fslwiki/) and ANTsPy (https://github.com/ANTsX/ANTsPy).

## Results

### Cohort description

A total of 1,022 patients from the INSPiRE-TMS cohort were included in the analysis. The mean age of the cohort was 66.3 years and 68.3% of the patients were male. Median NIHSS on admission was 2 (IQR 1-3), median mRS at baseline 1 (IQR 1-2) and median ischemic brain lesion volume 8.7 mL (IQR 2.7-33.9). Patient demographics as well as stroke-specific parameters were similar between the entire study population and the sub-group cohort used for subsequent LSM and LNM analyses (**Table 1**).

**Table 1.**
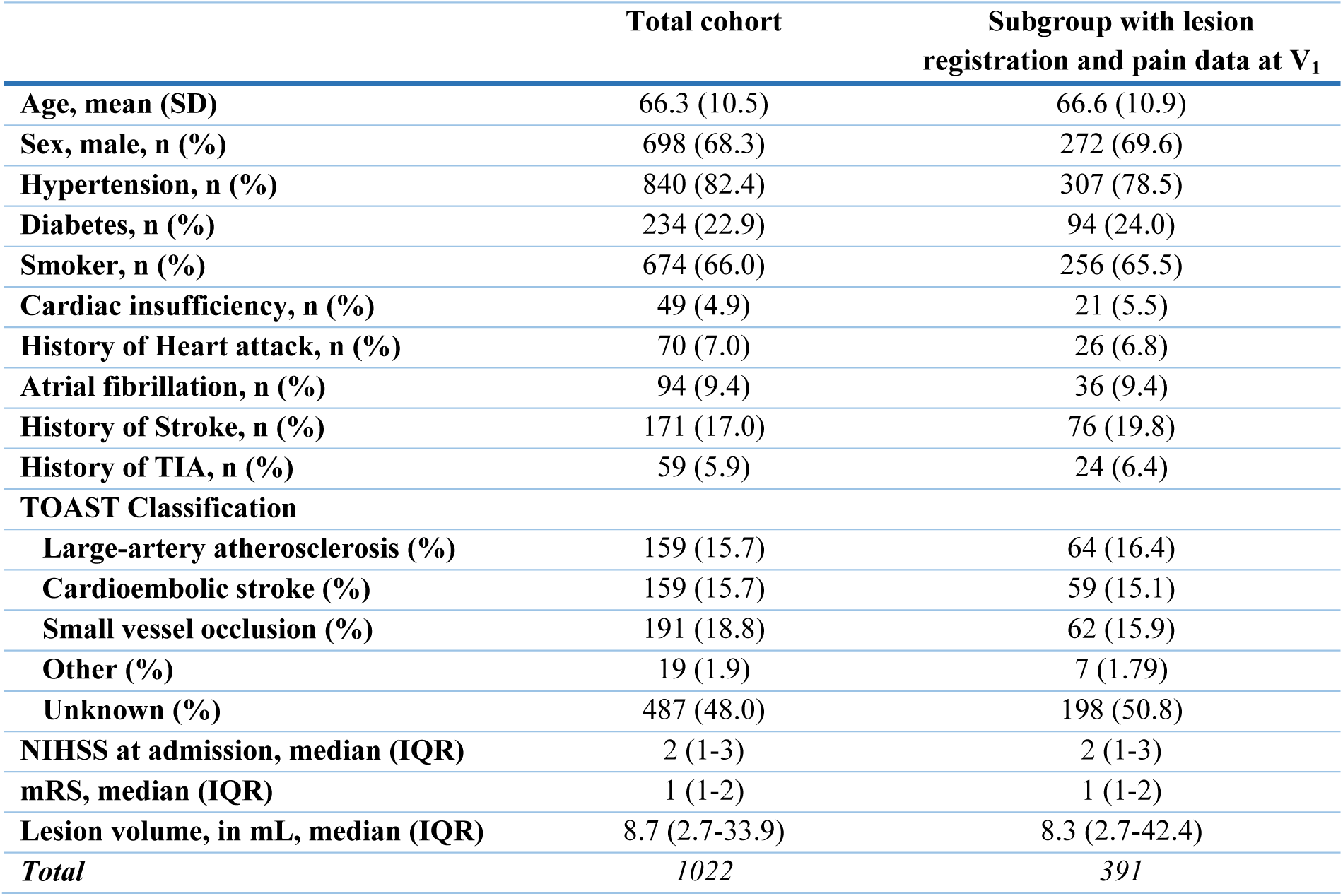
Cohort characteristics. mRS: modified Rankin Scale; NIHSS: National Institutes of Health Stroke Scale; TIA: transient ischemic attack; TOAST: Trial of Org 10172 in Acute Stroke Treatment.

|  | Total cohort | Subgroup with lesion registration and pain data at V<sub>1</sub> |
| --- | --- | --- |
| Age, mean (SD) | 66.3 (10.5) | 66.6 (10.9) |
| Sex, male, n (%) | 698 (68.3) | 272 (69.6) |
| Hypertension, n (%) | 840 (82.4) | 307 (78.5) |
| Diabetes, n (%) | 234 (22.9) | 94 (24.0) |
| Smoker, n (%) | 674 (66.0) | 256 (65.5) |
| Cardiac insufficiency, n (%) | 49 (4.9) | 21 (5.5) |
| History of Heart attack, n (%) | 70 (7.0) | 26 (6.8) |
| Atrial fibrillation, n (%) | 94 (9.4) | 36 (9.4) |
| History of Stroke, n (%) | 171 (17.0) | 76 (19.8) |
| History of TIA, n (%) | 59 (5.9) | 24 (6.4) |
| **TOAST Classification** |  |  |
| Large-artery atherosclerosis (%) | 159 (15.7) | 64 (16.4) |
| Cardioembolic stroke (%) | 159 (15.7) | 59 (15.1) |
| Small vessel occlusion (%) | 191 (18.8) | 62 (15.9) |
| Other (%) | 19 (1.9) | 7 (1.79) |
| Unknown (%) | 487 (48.0) | 198 (50.8) |
| NIHSS at admission, median (IQR) | 2 (1-3) | 2 (1-3) |
| mRS, median (IQR) | 1 (1-2) | 1 (1-2) |
| Lesion volume, in mL, median (IQR) | 8.7 (2.7-33.9) | 8.3 (2.7-42.4) |
| *Total* | *1022* | *391* |

Of the 1,022 patients included, 91% completed the EQ5D-3L pain dimension questionnaire at baseline (**Suppl. Table 1).** Pain (EQ5D-3L ≥ 2) was present in 43.1% at baseline (severe pain 3.1%) and increased by V_1_; about 5-6% reported severe pain at follow-up (**Suppl. Table 1**). The subgroup with imaging data available exhibited similar pain prevalence trends (**Suppl. Table 1**).

### Factors Associated with New Severe Pain

Of those 39 patients with severe pain at V_1_, 29 patients experienced “new severe pain” post-stroke, defined as patients who reported severe pain at V_1_ (EQ5D-3L = 3) but not at baseline (EQ5D-3L < 3 at baseline). In a two-group analysis comparing patients with “new severe pain” (N = 29) at V_1_ to those without “new severe pain” (N = 747), there was no statistically significant difference in terms of basic patient demographics (age, sex) or stroke-specific factors (cardiovascular risk factors, NIHSS, mRS, lesion volume, distribution of acute neurological deficits), quality of life measurements and depression scales (**Suppl. Table 2**).

Multivariable binary logistic regression identified baseline anxiety (assessed as 1, 2 or 3 in the EQ5D-3L questionnaire for anxiety) as being associated with “new severe pain” at V_1_ (odds ratio 2.9, 95% CI 1.17 – 7.17, p = 0.021). The model included age, male sex, baseline lesion volume, NIHSS and EQ5D-3l mobility as well; all these factors showed a weaker association with “new severe pain” at V_1_ than anxiety (**Table 2**).

**Table 2.** Multivariable binary logistic regression analysis for “new severe pain” at V_1_. 776 patients included, 29 having “new severe pain” at V_1_. Number of observations in multivariable model = 270. EQ5D-3L: European Quality of Life 5 Dimensions 3 Level Version.

| Multivariable binary logistic regression analysis for “new severe pain” (3) at V<sub>1</sub> |  |  |  |  |  |
| --- | --- | --- | --- | --- | --- |
|  | Bivariate model |  |  | Multivariable model |  |
|  | Odds ratio (95% CI) | P-value | Number of obs | Odds ratio (95% CI) | P-value |
| Age | 0.98 (0.95-1.01) | 0.29 | 776 | 0.99 (0.94-1.04) | 0.63 |
| Sex, Male | 0.74 (0.33-1.68) | 0.48 | 776 | 0.64 (0.17-2.46) | 0.52 |
| Lesion volume | 0.99 (0.92-1.07) | 0.83 | 291 | 0.99 (0.91-1.07) | 0.74 |
| NIHSS | 1.06 (0.94-1.20) | 0.35 | 776 | 0.95 (0.70-1.30) | 0.76 |
| mRS at Baseline | 1.32 (0.81-2.14) | 0.27 | 776 | 1.42 (0.57-3.56) | 0.45 |
| EQ5D-3L Anxiety at Baseline | 1.90 (1.00-3.58) | **0.048** | 721 | 2.90 (1.17-7.17) | **0.021** |
| EQ5D-3L Mobility at Baseline | 1.44 (0.66-3.14) | 0.35 | 721 | 2.83 (0.88-9.13) | 0.08 |

### Neuroimaging results

In a subgroup analysis of 391 patients for whom MRI data was available and could be used for LSM and LNM analyses, 18 (4.6%) reported “new severe pain” at V_1_. A heat map of all analyzed lesions of these patients is shown in **Figure 1A**. The LSM analysis for “new severe pain” at V_1_ did not reveal any voxels exceeding the significance threshold (p<0.05) in neither the voxel-wise nor atlas-based analyses. Voxel-wise LSM revealed weak and statistically non-significant associations with lesions in the occipital cortex, thalamus, and cerebellum, predominantly in the right hemisphere (**Figure 1B, Suppl. Table 3**).

**Figure 1.**
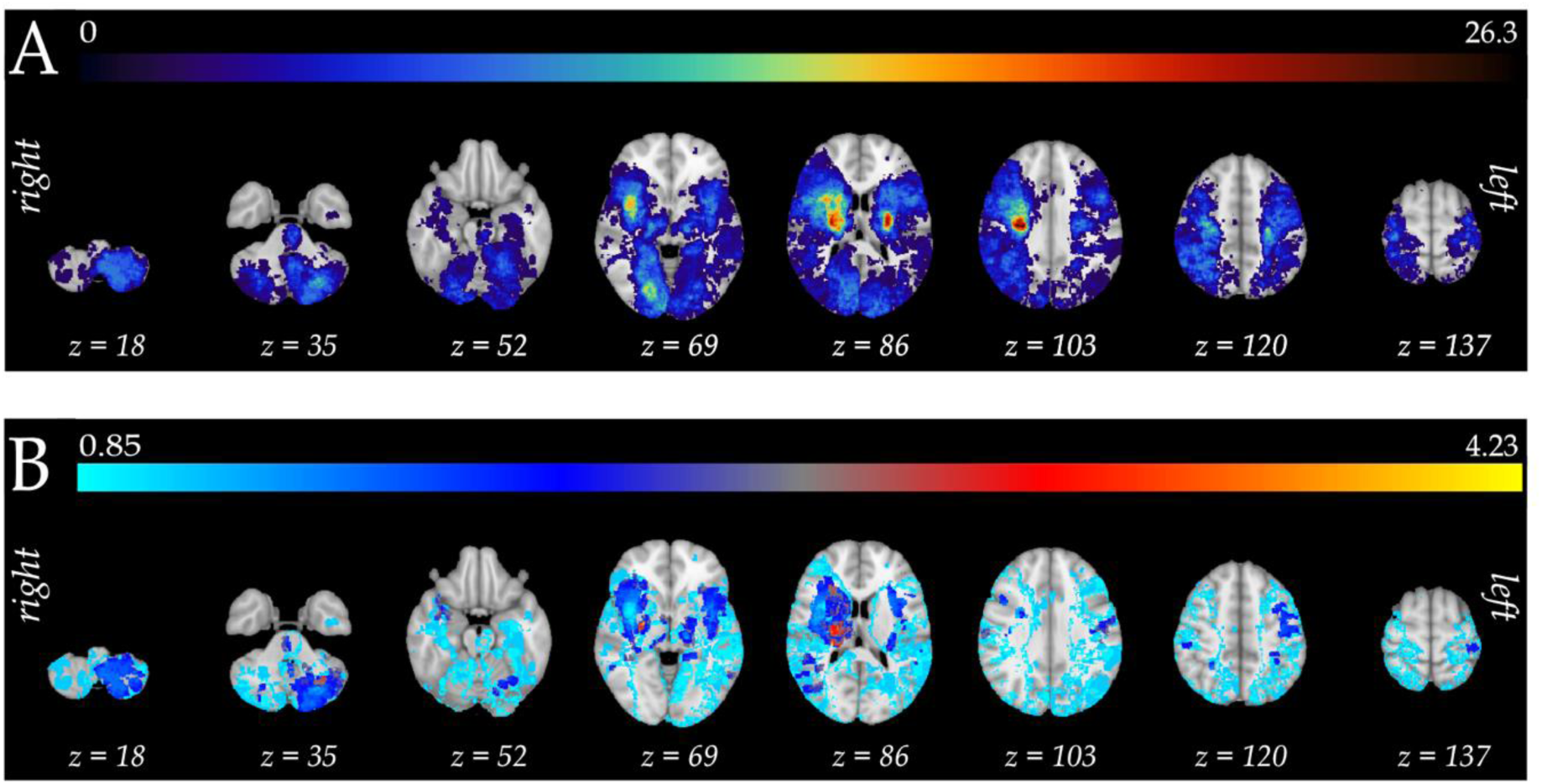
Lesion distribution and lesion symptom mapping for “new severe pain”. **A:** Lesion distribution map, overlap of all lesion maps (N = 391). Color scale indicates the number of patients showing a lesion in a given voxel; **B:** Results of LSM analysis relating lesion location to “new severe pain” at V_1_ (N = 18 patients); The color bar represents the strength of the statistical association of a lesion location with “new severe pain”. Lesion masks and results were overlaid on slices of an ex-vivo MNI space template. No voxels survived the significance threshold in LSM for “new severe pain”.

#### Lesion Network Mapping

The LNM analysis of “new severe pain” at V_1_ showed disruptions to a wide-spread bi-hemispheric network of regions including connections to cortical, subcortical, and cerebellar regions (**Figure 2A**, **C**). The network specifically included the frontal pole, cingulate gyrus, paracingulate gyrus, insular cortex, brainstem, thalamus, putamen, anterior thalamic radiation, and mainly crus I and VI of the cerebellum (**Table 3**). No regions were depicted using the significance threshold of pFWE<0.05. A sensitivity analysis adjusting for anxiety as a covariate revealed a network similar to “new severe pain” alone (**Figure 2B**, **Suppl. Figure 2A**), with highest connectivity values to insular cortex, frontal operculum cortex, thalamus, anterior thalamic radiation, crus VI and I of the cerebellum (**Suppl. Figure 2B**, **Table 3**). Regions that were depicted using the significance threshold of pFWE<0.05 included, among others, insular cortex, thalamus, putamen, pallidum, caudate, anterior thalamic radiation, inferior fronto-occipital fasciculus, corticospinal tract, and various cerebellar regions (**Table 3).** An additional sensitivity analysis of patients who have had severe pain at V_1_ (EQ5D-3L = 3) but did not report any pain at baseline (EQ5D-3L = 0) (N = 5 patients) was conducted. This analysis showed a similar network (**Suppl. Figure 2C**) to the “new severe pain” network (**Figure 2C**), with overlapping regions particularly in cingulate cortex and insular cortex.

**Figure 2.**
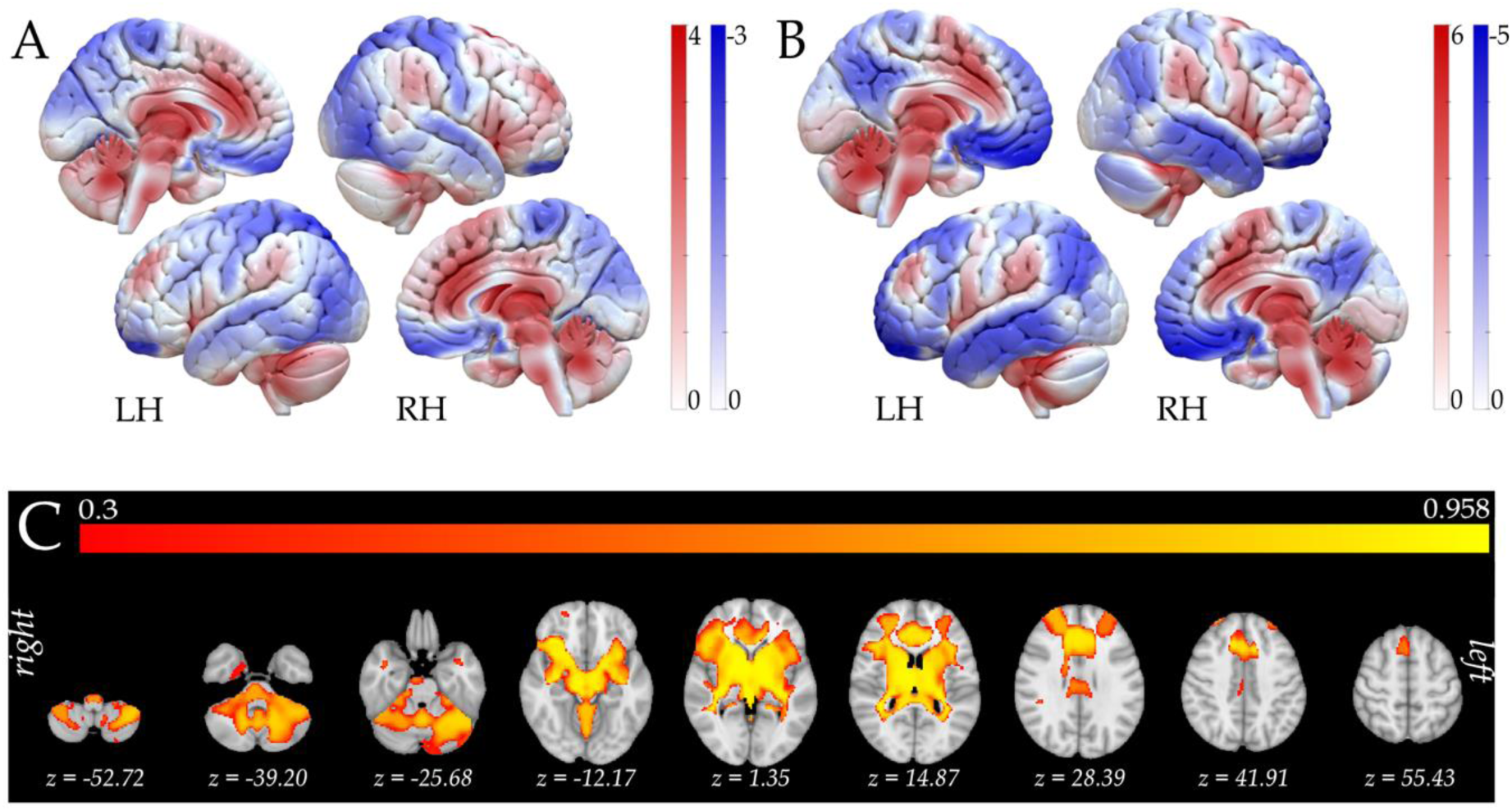
Lesion network mapping for “new severe pain”. **A:** Maps derived from permutation analysis using “new severe pain” (N = 18 patients) as the variable of interest overlaid on a 3D surface of the brain, positive correlations shown in red, negative correlations shown in blue; **B:** LNM result for “new severe pain” at V_1_ with anxiety as a covariate (N = 18 patients); **C:** Voxels associated with “new severe pain” at V_1_ (N = 18 patients). FWE-corrected map thresholded at p<0.05 (1-p shown). LH indicates left hemisphere; and RH, right hemisphere.

**Table 3.**
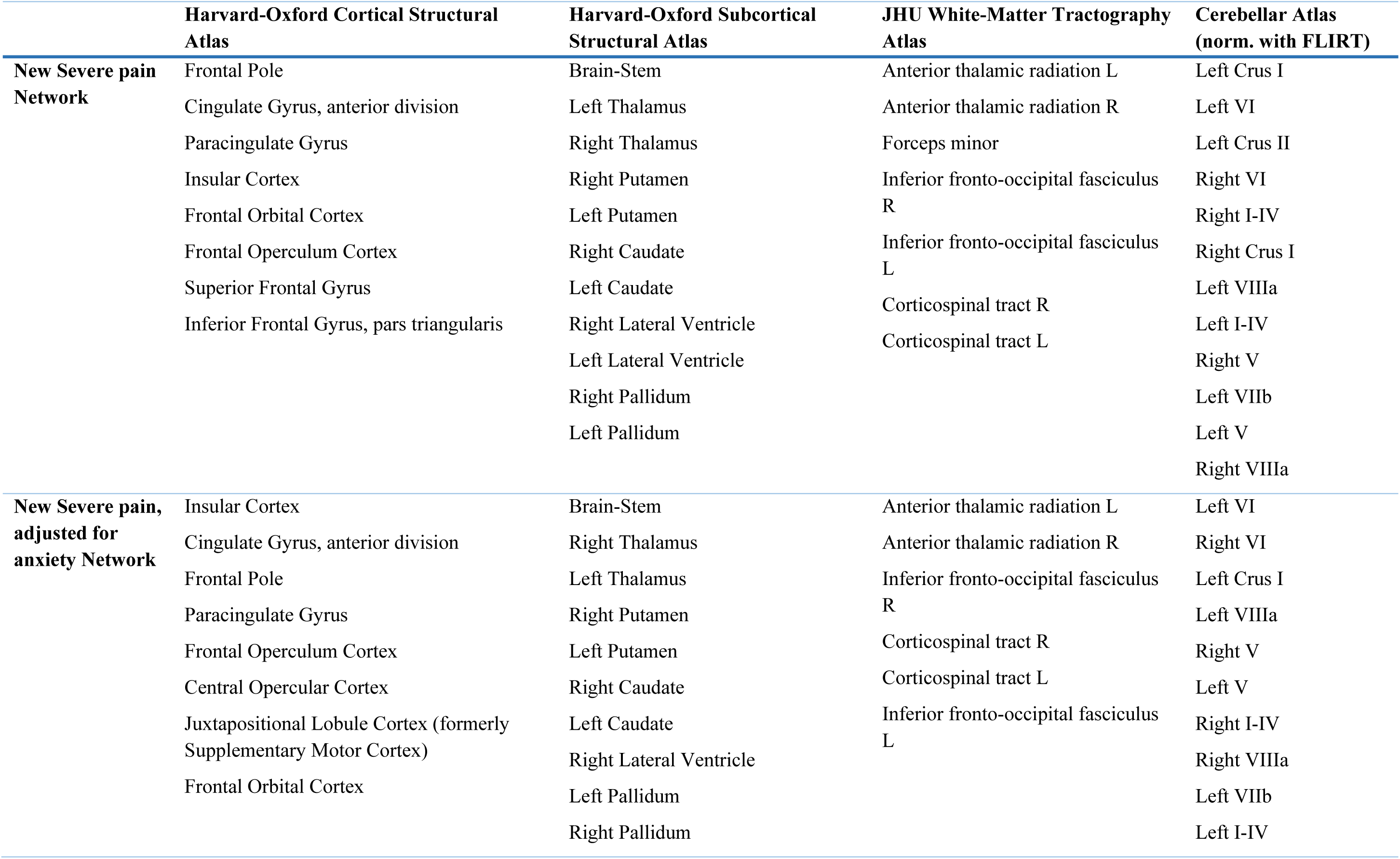
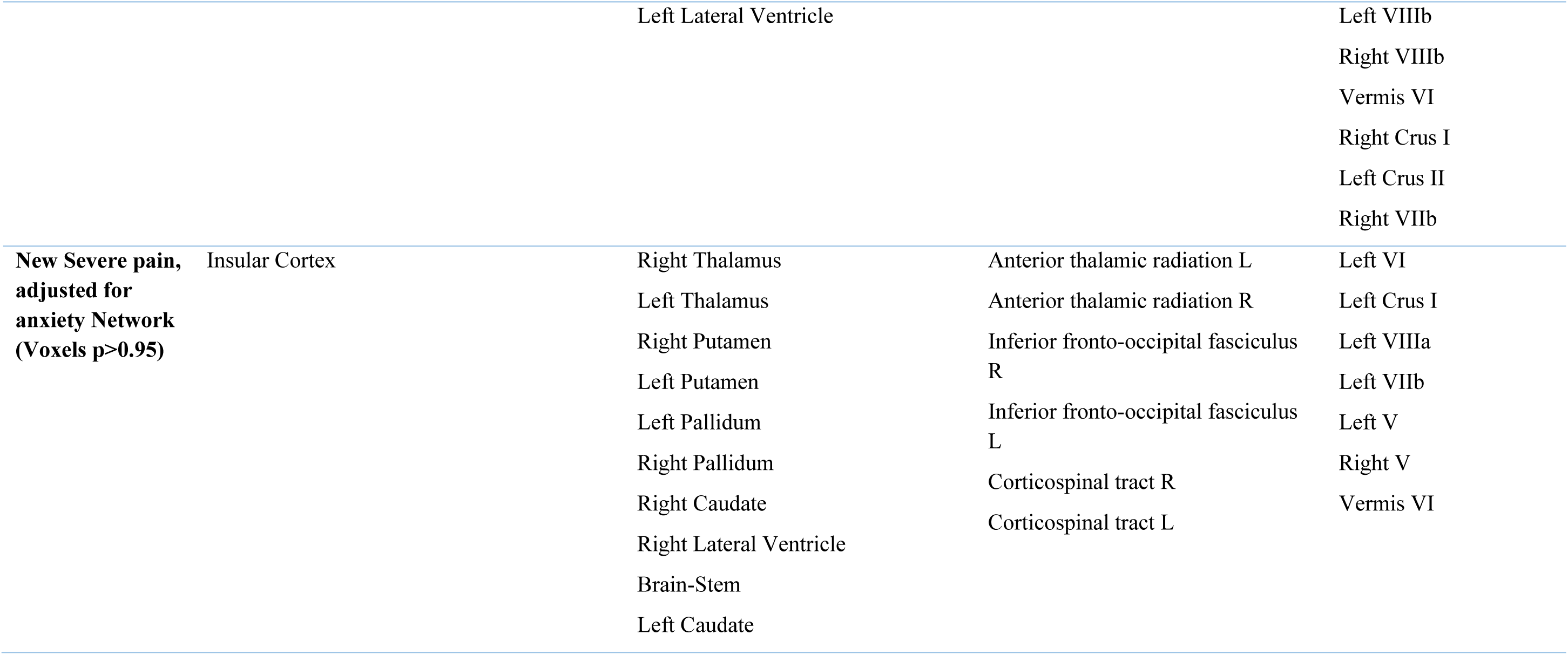
Atlas based labelling of lesion network mapping results anatomical description.

#### Comparison to Neurosynth

The decoding tool available on the Neurosynth platform which encompasses data from 14,371 studies (https://www.neurosynth.org) was used to systematically compare the spatial similarity of our identified networks with all the maps in its database.

Out of 1,334 anatomical and functional terms, Neurosynth comparisons revealed that the top 25 similarities to the unthresholded “new severe pain with anxiety as covariate” map (**Figure 2B**) included terms specifically associated with “*pain*”, “*painful*”, “*noxious*” and “*nociceptive*” (**Figure 3A, B**). In contrast, the “new severe pain” showed top 25 similarities exclusively related to anatomical terms (**Suppl. Table 4**).

**Figure 3.**
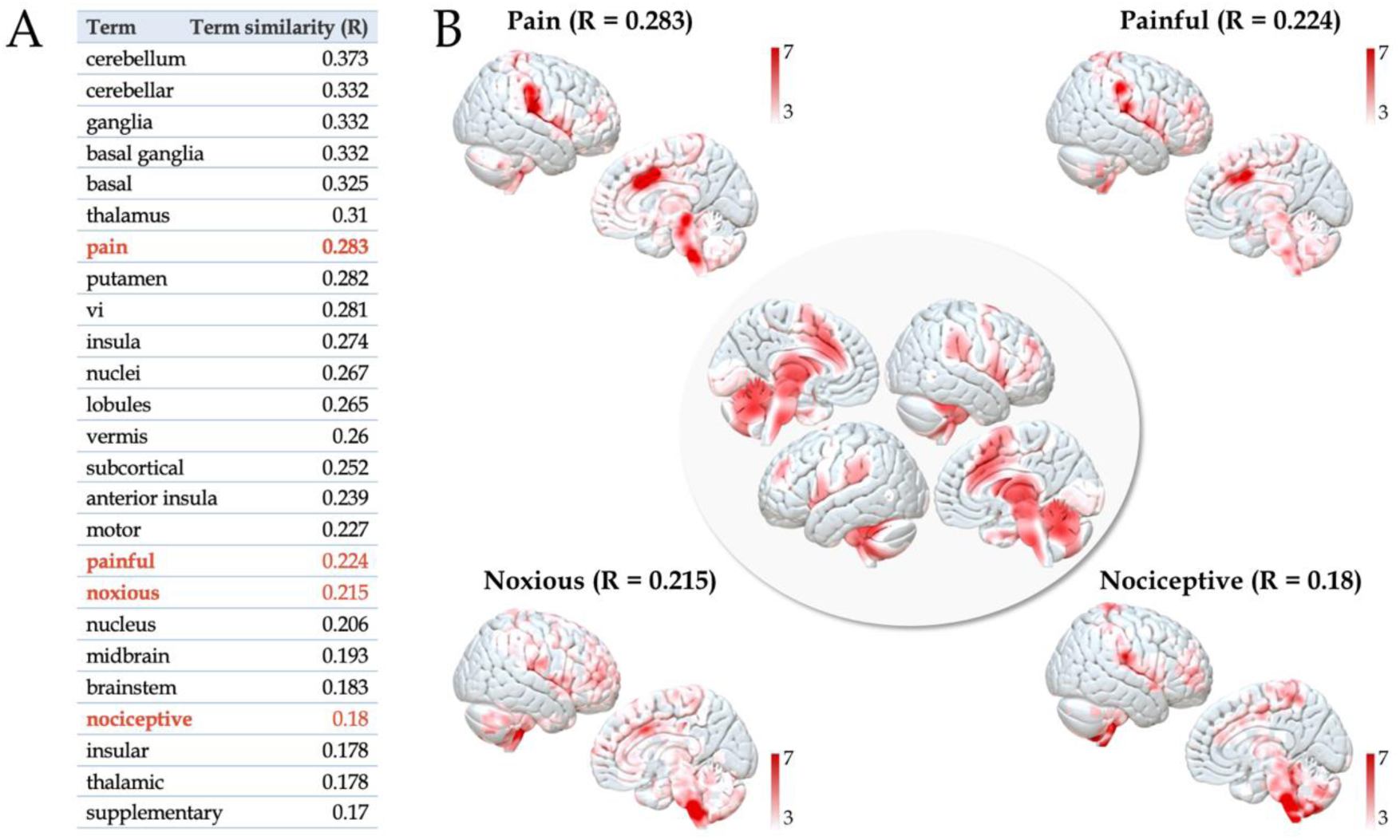
“New severe pain” network compared to association maps from Neurosynth. **A:** Top 25 associated terms to “new severe pain” (with anxiety as a covariate), purely anatomical terms are shown in black, non-anatomical terms shown are in red; **B:** In the middle depiction of LNM results of “new severe pain” with anxiety as covariate, around it association test maps of non-anatomical terms from Neurosynth decoder results overlaid on a brain surface. Color bar indicates T-score.

## Discussion

This study provides new insights into the prevalence and characteristics of post stroke pain (PSP) and the neural correlates of PSP, highlighting the complex interplay between psychological factors, lesion characteristics and brain network dysfunction. These findings contribute to a growing understanding of PSP as a complex condition requiring a multidisciplinary approach to treatment and rehabilitation.

In our cohort, up to 50% of stroke survivors reported any PSP, with severe pain affecting 5-6% of patients. These findings align with previous studies^2,3,7^, which observed similar pain prevalence despite differences in stroke severity. Notably, the mild median NIHSS score in our cohort emphasizes that PSP is not solely a consequence of severe strokes. Our results further suggest an association between psychological factors and pain outcomes, identifying anxiety as the main characteristic associated with new severe PSP. This is consistent with the well-documented role of psychological factors in different chronic pain conditions^10–12^, but has not been described in PSP so far. Notably, Naess et al.^33^ reported pain, depression and fatigue as a symptom-cluster arising after stroke. These findings underscore the need for early psychological assessment and intervention. Unaddressed psychological comorbidities may exacerbate PSP, impede recovery, and contribute to adverse outcomes, as chronic pain itself is associated with higher 10-year mortality^34^.

Neuroimaging analyses were performed on a subgroup closely resembling the total cohort in terms of patient-specific characteristics, including a mild median NIHSS. Although voxel- and atlas-based LSM analyses did not yield statistically significant results, they identified weak and statistically not significant associations between severe pain and lesions in the occipital cortex, thalamus, and cerebellum. This aligns with prior evidence linking these regions to PSP^14,15^.

However, the notion of a single ”pain region” in the brain is apparently overly simplistic. Pain perception is a complex cognitive process involving multiple regions and networks, often referred to as the “neurological signature of physical pain”^35^, with significant inter-subject variability influenced by previous pain experiences and psychological states^10^.

Despite the generally mild stroke severity of patients included in our study with corresponding small ischemic brain lesion sizes, LNM revealed a coherent network associated with PSP, identifying disruptions in a distributed network encompassing the anterior cingulate cortex (ACC), insular cortex and thalamus. Adjusting our LNM analyses for anxiety refined the pain network further, underscoring the interactive effects of psychological states on pain perception.

Notably, the insular cortex and ACC play a central role in the affective and valence-related dimensions of chronic pain. The insular cortex is implicated in interoceptive awareness^36,37^, while the ACC is associated with pain’s aversive and cognitive components^38,39^ and is an established target in neuromodulatory pain therapies^40,41^. Being involved in the limbic system, it may be particularly effective as a therapeutic target in pain patients with affective comorbidities, though further research is warranted. The thalamus, a structurally and functionally multifaceted region, acts as a key hub in our severe pain network. Its ventral posterolateral (VPL) and ventral posteromedial (VPM) nuclei transmit nociceptive information to the somatosensory cortex, influencing both the sensory-discriminative aspects of pain perception^42,43^, and pain intensity itself^44^. Additionally, the intralaminar nuclei and mediodorsal nucleus, connected to limbic structures such as the ACC^43^ and the primary motor cortex^45^, further modulate the affective-motivational and motor components of pain^43^. The involvement in pain processing is supported by clinical studies demonstrating that thalamic neuromodulation, specifically targeting the VPM/VPL nuclei, has been used therapeutically to alleviate chronic pain^41,46^.

Overall, these findings support the notion that PSP is linked not to distinct lesion locations but rather to disruptions within a functionally connected defined network, reinforcing the view that neuromodulatory pain therapy should target networks rather than single regions^41^.

Although we could not differentiate for pain etiology (vide infra) – given the limited information on pain character and onset – our results overlap with those reported by Elias et al.^14^, who investigated solely central neuropathic PSP. Their findings similarly implicated the insular cortex, thalamus and ACC in PSP, reinforcing the robustness of our results and emphasizing the strong interaction with specific disrupted networks and pain perception independent of pain etiology. Neurosynth comparisons support this notion, as our *pain+anxiety* network most strongly overlapped with pain-related terms that were defined meta analytically and without etiologic context.

Approximately two-thirds of patients with central PSP do not receive adequate pain therapy^47^, despite evidence that effective pain treatment is associated with improvements in cognition and quality of life^9^. Although the median NIHSS was low, the high prevalence of severe pain in our cohort highlights the importance of assessing and treating pain even in mild ischemic strokes. The strong association between PSP and anxiety highlights the need to investigate a multidisciplinary approach to pain management. Traditional treatments focusing solely on somatic pain mechanisms may be insufficient, particularly in patients with psychological comorbidities. Moreover, longitudinal studies are needed to evaluate how optimized pain therapy may affect long-term outcomes and stroke recurrence. Understanding the bidirectional relationship between pain and psychological states could further refine treatment protocols and improve quality of life for stroke survivors.

Neuromodulation may offer additional relief, considering our findings of network disruptions associated with severe pain. LNM findings could help define both invasive and non-invasive neuromodulation targets for PSP patients with or without anxiety – a strategy that has proven successful for other symptoms^48,49^. Further studies are now needed to validate these network targets.

Key strengths of the present study include its large, well-characterized cohort stemming from a randomized controlled trial and the integration of both clinical and neuroimaging data. The use of LNM provided a network-level perspective on PSP, offering novel insights into its neural underpinnings. By including patients with heterogenous lesion locations – both cortical and subcortical – we minimized bias related to lesion site, a strength compared to other pain-LNM studies that, for example, focused specifically on thalamic lesions^15^.

## Limitations

The most important limitation is the study’s retrospective nature, which provided only limited information on pain quality, etiology and potential pre-existing pain syndromes. We were hence unable to make inferences about pain etiology in our cohort. Future prospective studies should address these important gaps by incorporating comprehensive assessments of pain characteristics, pre-stroke pain history, pain medication usage, as well as the impact of comorbid conditions, such as depression and anxiety disorders, on PSP. Furthermore, the use of LNM with normative connectomes provides an indirect measure of functional disruption, hence explaining symptoms with reduced variance compared to direct measures of functional connectivity. Additionally, for this type of LNM analysis, averaging of BOLD signals within the entire lesion mask is required, which can be problematic specifically in large lesions spanning grey and white matter. Prospective studies are needed to identify the specific contributions of grey and white matter in the development of pain after stroke^50^. Nonetheless, this type of methodology is advantageous by facilitating the analysis of large amounts of lesion data from patients coming directly from a clinical setting in which functional MRI scanning is not part of the patient care routine, allowing us to identify what the lesioned area would be connected to in the healthy brain, potentially shedding light into causal disruptions associated with the presence of a specific symptom.

## Conclusion

PSP is a significant burden for stroke survivors. It likely arises from a complex interplay of lesion location, network dysfunction, and psychosocial factors. This study has the following major findings: (1) up to 50% of stroke survivors experience any PSP, (2) anxiety emerges as a critical associated factor for new-onset severe pain, and (3) neuroimaging reveals disruptions in a key brain network associated with PSP, including the ACC and insular cortex, providing a neuroanatomical substrate of PSP. These findings underscore the need for integrated, multidisciplinary care to address the multifaceted nature of PSP. By leveraging insights from network-level analyses, clinicians can move toward more personalized and effective treatment strategies, ultimately improving long-term outcomes and quality of life.

## Author contributions

AS and AKu designed the study and prepared the manuscript. ASR delineated lesions., and AKu, AKh, RG and BB supervised lesion delineation. AS, ASR and TU performed the pre-processing of the imaging data. AS performed regression analysis on demographic and clinical data, and ran LSM and LNM analyses. ASR, AKh, UG, TU, AH, HA and ME participated in study design and interpretation of results. AKu and ME jointly supervised this work.

## Data Availability

Data supporting the results of this study is available upon request from the corresponding author.

## Acknowledgement

We would like to thank all participants in this study for their invaluable contribution. Additionally, we thank the INSPiRE-TMS team for collecting and providing the data. A.H. was supported by the German Research Foundation (Deutsche Forschungsgemeinschaft, 424778381 – TRR 295), Deutsches Zentrum für Luft- und Raumfahrt (DynaSti grant within the EU Joint Programme Neurodegenerative Disease Research, JPND), the National Institutes of Health (R01MH130666, 1R01NS127892-01, 2R01 MH113929 & UM1NS132358) as well as the New Venture Fund (FFOR Seed Grant).

## Supplemental Material

Suppl. Tables 1-4

Suppl. Figures 1-2

## Non-standard Abbreviations and Acronyms

ACC: anterior cingulate cortex
BOLD: blood-Oxygenation-Level Dependent
DWI: diffusion-weighted imaging
EQ5D-3L: European Quality of Life 5 Dimensions 3 Level Version
FLAIR: fluid-attenuated inversion recovery
FWE: family wise error
INSPiRE-TMS: Intensified secondary prevention intending a reduction of recurrent events in TIA and minor stroke patients
LNM: lesion-network mapping
LSM: lesion-symptom mapping
MRI: magnetic resonance imaging
mRS: modified Rankin Scale
NIHSS: National Institutes of Health Stroke Scale
PSP: post-stroke pain
rs-fMRI: resting-state functional magnetic resonance imaging
TFCE: threshold-free cluster enhancement
TIA: transient ischemic attack
TOAST: Trial of Org 10172 in Acute Stroke Treatment
VPL: ventral posterolateral
VPM: ventral posteromedial

## References

1. Westerlind E, Singh R, Persson HC, Sunnerhagen KS. Experienced pain after stroke: a cross-sectional 5-year follow-up study. BMC Neurol. 2020;20:4. doi: 10.1186/s12883-019-1584-z

2. Ali M, Tibble H, Brady MC, Quinn TJ, Sunnerhagen KS, Venketasubramanian N, Shuaib A, Pandyan A, Mead G, Lees KR, et al. Prevalence, Trajectory, and Predictors of Poststroke Pain: Retrospective Analysis of Pooled Clinical Trial Data Set. Stroke. 2023;54:3107–3116. doi: 10.1161/strokeaha.123.043355

3. Hansen AP, Marcussen NS, Klit H, Andersen G, Finnerup NB, Jensen TS. Pain following stroke: a prospective study. Eur J Pain. 2012;16:1128–1136. doi: 10.1002/j.1532-2149.2012.00123.x

4. Lundstrom E, Smits A, Terent A, Borg J. Risk factors for stroke-related pain 1 year after first-ever stroke. Eur J Neurol. 2009;16:188–193. doi: 10.1111/j.1468-1331.2008.02378.x

5. Payton H, Soundy A. The Experience of Post-Stroke Pain and The Impact on Quality of Life: An Integrative Review. Behav Sci (Basel). 2020;10. doi: 10.3390/bs10080128

6. Hoang CL, Salle JY, Mandigout S, Hamonet J, Macian-Montoro F, Daviet JC. Physical factors associated with fatigue after stroke: an exploratory study. Top Stroke Rehabil. 2012;19:369–376. doi: 10.1310/tsr1905-369

7. Naess H, Lunde L, J B. The effects of fatigue, pain, and depression on quality of life in ischemic stroke patients: The Bergen Stroke Study. Vasc Health Risk Manag. 2012;8:407–413. doi: 10.2147/VHRM.S32780

8. O’Donnell MJ, Diener HC, Sacco RL, Panju AA, Vinisko R, Yusuf S, Investigators PR. Chronic pain syndromes after ischemic stroke: PRoFESS trial. Stroke. 2013;44:1238–1243. doi: 10.1161/STROKEAHA.111.671008

9. Harrison RA, Field TS. Post Stroke Pain: Identification, Assessment, and Therapy. Cerebrovascular Diseases. 2015;39:190–201. doi: 10.1159/000375397

10. Nees F, Becker S. Psychological Processes in Chronic Pain: Influences of Reward and Fear Learning as Key Mechanisms - Behavioral Evidence, Neural Circuits, and Maladaptive Changes. Neuroscience. 2018;387:72–84. doi: 10.1016/j.neuroscience.2017.08.051

11. Rogers AH, Farris SG. A meta-analysis of the associations of elements of the fear-avoidance model of chronic pain with negative affect, depression, anxiety, pain-related disability and pain intensity. Eur J Pain. 2022;26:1611–1635. doi: 10.1002/ejp.1994

12. Turk DC, Okifuji A. Psychological factors in chronic pain: evolution and revolution. J Consult Clin Psychol. 2002;70:678–690. doi: 10.1037//0022-006x.70.3.678

13. Hong JH, Bai DS, Jeong JY, Choi BY, Chang CH, Kim SH, Ahn SH, Jang SH. Injury of the spino-thalamo-cortical pathway is necessary for central post-stroke pain. Eur Neurol. 2010;64:163–168. doi: 10.1159/000319040

14. Elias GJB, De Vloo P, Germann J, Boutet A, Gramer RM, Joel SE, Morlion B, Nuttin B, Lozano AM. Mapping the network underpinnings of central poststroke pain and analgesic neuromodulation. Pain. 2020;161:2805–2819. doi: 10.1097/j.pain.0000000000001998

15. Kim NY, Taylor JJ, Kim YW, Borsook D, Joutsa J, Li J, Quesada C, Peyron R, Fox MD. Network Effects of Brain Lesions Causing Central Poststroke Pain. Annals of Neurology. 2022;92:834–845. doi: 10.1002/ana.26468

16. Klit H, Finnerup NB, Andersen G, Jensen TS. Central poststroke pain: a population-based study. Pain. 2011;152:818–824. doi: 10.1016/j.pain.2010.12.030

17. Krause T, Asseyer S, Taskin B, Floel A, Witte AV, Mueller K, Fiebach JB, Villringer K, Villringer A, Jungehulsing GJ. The Cortical Signature of Central Poststroke Pain: Gray Matter Decreases in Somatosensory, Insular, and Prefrontal Cortices. Cereb Cortex. 2016;26:80–88. doi: 10.1093/cercor/bhu177

18. Liampas A, Velidakis N, Georgiou T, Vadalouca A, Varrassi G, Hadjigeorgiou GM, Tsivgoulis G, Zis P. Prevalence and Management Challenges in Central Post-Stroke Neuropathic Pain: A Systematic Review and Meta-analysis. Adv Ther. 2020;37:3278–3291. doi: 10.1007/s12325-020-01388-w

19. Kumar G, Soni CR. Central post-stroke pain: current evidence. J Neurol Sci. 2009;284:10–17. doi: 10.1016/j.jns.2009.04.030

20. Convers P, Creac’h C, Beschet A, Laurent B, Garcia-Larrea L, Peyron R. A hidden mesencephalic variant of central pain. Eur J Pain. 2020;24:1393–1399. doi: 10.1002/ejp.1588

21. Fox MD. Mapping Symptoms to Brain Networks with the Human Connectome. N Engl J Med. 2018;379:2237–2245. doi: 10.1056/NEJMra1706158

22. Boes AD, Prasad S, Liu H, Liu Q, Pascual-Leone A, Caviness VS, Jr., Fox MD. Network localization of neurological symptoms from focal brain lesions. Brain. 2015;138:3061–3075. doi: 10.1093/brain/awv228

23. Ahmadi M, Laumeier I, Ihl T, Steinicke M, Ferse C, Endres M, Grau A, Hastrup S, Poppert H, Palm F, et al. A support programme for secondary prevention in patients with transient ischaemic attack and minor stroke (INSPiRE-TMS): an open-label, randomised controlled trial. Lancet Neurol. 2020;19:49–60. doi: 10.1016/S1474-4422(19)30369-2

24. Leistner S, Michelson G, Laumeier I, Ahmadi M, Smyth M, Nieweler G, Doehner W, Sobesky J, Fiebach JB, Marx P, et al. Intensified secondary prevention intending a reduction of recurrent events in TIA and minor stroke patients (INSPiRE-TMS): a protocol for a randomised controlled trial. BMC Neurology 2013.

25. Rangus I, Rios AS, Horn A, Fritsch M, Khalil A, Villringer K, Udke B, Ihrke M, Grittner U, Galinovic I, et al. Fronto-thalamic networks and the left ventral thalamic nuclei play a key role in aphasia after thalamic stroke. Commun Biol. 2024;7:700. doi: 10.1038/s42003-024-06399-9

26. Jenkinson M, Beckmann C, Behrens T, Woolrich M, Smith S. FSL. Neuroimage. 2012;62. doi: 10.1016/j.neuroimage.2011.09.015

27. Fonov VS, Evans, A. C., McKinstry, R. C., Almli, C. R., & Collins, D. L. Unbiased nonlinear average age-appropriate brain templates from birth to adulthood. NeuroImage. 2009;47.

28. Avants BB, Tustison NJ, Song G, Cook PA, Klein A, Gee JC. A reproducible evaluation of ANTs similarity metric performance in brain image registration. Neuroimage. 2011;54:2033–2044. doi: 10.1016/j.neuroimage.2010.09.025

29. DeMarco AT, Turkeltaub PE. A multivariate lesion symptom mapping toolbox and examination of lesion-volume biases and correction methods in lesion-symptom mapping. Hum Brain Mapp. 2018;39:4169–4182. doi: 10.1002/hbm.24289

30. Lugtmeijer S, Geerligs L, de Leeuw FE, de Haan EHF, Kessels RPC, Visual Brain G. Are visual working memory and episodic memory distinct processes? Insight from stroke patients by lesion-symptom mapping. Brain Struct Funct. 2021;226:1713–1726. doi: 10.1007/s00429-021-02281-0

31. Holmes AJ, Hollinshead MO, O’Keefe TM, Petrov VI, Fariello GR, Wald LL, Fischl B, Rosen BR, Mair RW, Roffman JL, et al. Brain Genomics Superstruct Project initial data release with structural, functional, and behavioral measures. Sci Data. 2015;2:150031. doi: 10.1038/sdata.2015.31

32. Gorgolewski KJ, Varoquaux G, Rivera G, Schwartz Y, Sochat VV, Ghosh SS, Maumet C, Nichols TE, Poline JB, Yarkoni T, et al. NeuroVault.org: A repository for sharing unthresholded statistical maps, parcellations, and atlases of the human brain. Neuroimage. 2016;124:1242–1244. doi: 10.1016/j.neuroimage.2015.04.016

33. Naess H, Lunde L, Brogger J. The triad of pain, fatigue and depression in ischemic stroke patients: the Bergen Stroke Study. Cerebrovasc Dis. 2012;33:461–465. doi: 10.1159/000336760

34. Torrance N, Elliott AM, Lee AJ, Smith BH. Severe chronic pain is associated with increased 10 year mortality. A cohort record linkage study. Eur J Pain. 2010;14:380–386. doi: 10.1016/j.ejpain.2009.07.006

35. Wager TD, Atlas LY, Lindquist MA, Roy M, Woo CW, Kross E. An fMRI-based neurologic signature of physical pain. N Engl J Med. 2013;368:1388–1397. doi: 10.1056/NEJMoa1204471

36. Labrakakis C. The Role of the Insular Cortex in Pain. Int J Mol Sci. 2023;24. doi: 10.3390/ijms24065736

37. Lu C, Yang T, Zhao H, Zhang M, Meng F, Fu H, Xie Y, Xu H. Insular Cortex is Critical for the Perception, Modulation, and Chronification of Pain. Neurosci Bull. 2016;32:191–201. doi: 10.1007/s12264-016-0016-y

38. Fuchs PN, Peng YB, Boyette-Davis JA, Uhelski ML. The anterior cingulate cortex and pain processing. Front Integr Neurosci. 2014;8:35. doi: 10.3389/fnint.2014.00035

39. Xiao X, Zhang YQ. A new perspective on the anterior cingulate cortex and affective pain. Neurosci Biobehav Rev. 2018;90:200–211. doi: 10.1016/j.neubiorev.2018.03.022

40. Xiao X, Ding M, Zhang YQ. Role of the Anterior Cingulate Cortex in Translational Pain Research. Neurosci Bull. 2021;37:405–422. doi: 10.1007/s12264-020-00615-2

41. Motzkin JC, Kanungo I, D’Esposito M, Shirvalkar P. Network targets for therapeutic brain stimulation: towards personalized therapy for pain. Front Pain Res (Lausanne). 2023;4:1156108. doi: 10.3389/fpain.2023.1156108

42. Cao B, Xu Q, Shi Y, Zhao R, Li H, Zheng J, Liu F, Wan Y, Wei B. Pathology of pain and its implications for therapeutic interventions. Signal Transduct Target Ther. 2024;9:155. doi: 10.1038/s41392-024-01845-w

43. Kuner R, Kuner T. Cellular Circuits in the Brain and Their Modulation in Acute and Chronic Pain. Physiol Rev. 2021;101:213–258. doi: 10.1152/physrev.00040.2019

44. You HJ, Lei J, Pertovaara A. Thalamus: The ’promoter’ of endogenous modulation of pain and potential therapeutic target in pathological pain. Neurosci Biobehav Rev. 2022;139:104745. doi: 10.1016/j.neubiorev.2022.104745

45. Gan Z, Gangadharan V, Liu S, Korber C, Tan LL, Li H, Oswald MJ, Kang J, Martin-Cortecero J, Mannich D, et al. Layer-specific pain relief pathways originating from primary motor cortex. Science. 2022;378:1336–1343. doi: 10.1126/science.add4391

46. Tan H, Yamamoto EA, Elkholy MA, Raslan AM. Treating Chronic Pain with Deep Brain Stimulation. Curr Pain Headache Rep. 2023;27:11–17. doi: 10.1007/s11916-022-01099-7

47. Haslam BS, Butler DS, Kim AS, Carey LM. Chronic pain following stroke: Current treatment and perceived effect. Disability and Health Journal. 2021;14. doi: 10.1016/j.dhjo.2020.100971

48. Weigand A, Horn A, Caballero R, Cooke D, Stern AP, Taylor SF, Press D, Pascual-Leone A, Fox MD. Prospective Validation That Subgenual Connectivity Predicts Antidepressant Efficacy of Transcranial Magnetic Stimulation Sites. Biol Psychiatry. 2018;84:28–37. doi: 10.1016/j.biopsych.2017.10.028

49. Joutsa J, Shih LC, Horn A, Reich MM, Wu O, Rost NS, Fox MD. Identifying therapeutic targets from spontaneous beneficial brain lesions. Ann Neurol. 2018;84:153–157. doi: 10.1002/ana.25285

50. Salvalaggio A, Pini L, De Filippo De Grazia M, Thiebaut De Schotten M, Zorzi M, Corbetta M. Reply: Lesion network mapping: where do we go from here? Brain. 2021;144:e6. doi: 10.1093/brain/awaa351

